# Pilot Study Demonstrates Benefits of Nursing Home Air Purification on COVID-19 Outcomes

**DOI:** 10.1101/2022.12.01.22282978

**Authors:** Eric Jutkowitz, Peter Shewmaker, Ann Reddy, Joseph M. Braun, Rosa R. Baier

**Affiliations:** Department of Health Services, Policy & Practice, Brown University School of Public Health, Providence, RI; Evidence Synthesis Program Center Providence VA Medical Center, Providence, RI; Center of Innovation in Long-Term Services and Supports, Providence VA Medical Center, Providence, RI; Center for Long-Term Care Quality & Innovation, Brown University School of Public Health, Providence, RI; Department of Epidemiology, Brown University School of Public Health, Providence, RI

**Keywords:** nursing homes, SARS-CoV-2, COVID-19, indoor air, air purification, ultraviolet

## Abstract

Improving indoor air quality is one potential strategy to reduce the transmission of SARS-CoV-2 in any setting, including nursing homes, where staff and residents have been disproportionately and negatively affected by the COVID-19 pandemic. We used an interrupted time series design to compare trends in weekly COVID-19 cases and deaths before and after installation of ultraviolet air purification in 84 nursing homes in Florida, Georgia, North Carolina, and South Carolina from September 31, 2020 to December 27, 2020. Compared to pre-installation, weekly COVID-19 cases per 1,000 residents (−1.69, 95%CI: -4.32, 0.95) and the weekly probability of reporting any COVID-19 case (−0.02, 95%CI: -0.04, 0.00) declined in the post-installation period. We did not find any difference pre- and post-installation in COVID-19-related mortality (0.00 95%CI: -0.01, 0.02). Our findings from this small number of nursing homes in the southern US demonstrate the potential benefits of air purification in nursing homes on COVID-19 outcomes. We recommend a stronger experimental design to estimate the causal effect of installing air purification devices like this one on improving COVID-19 outcomes in nursing homes.

**Practical Implications:** Improving indoor air quality is one potential strategy to reduce the burden of COVID-19 in nursing homes and nursing homes are eligible to receive Civil Monetary Penalty funding for purchases that improve air quality. Intervening on air quality may have a wide impact without placing significant burden on individuals to modify their behavior. In this pilot evaluation, we found that installing ultraviolet air purification may be an effective strategy to reduce COVID-19 cases in nursing homes. We recommend a stronger experimental design to determine the causal effect of indoor air interventions, such as air purification, on COVID-19 in this setting.

## Introduction

In the United States (US), the COVID-19 pandemic has disproportionately affected both staff and residents in nursing homes.^1-4^ COVID-19 mortality among residents, specifically, has fluctuated over the course of the pandemic, but has, at times, accounted for more than 50% of all US COVID-19 deaths.^2^ Residents are vulnerable to COVID-19 for several reasons. First, nursing homes have many staff and visitors (medical professionals and family/friends) who frequently have direct contact with residents and are the primary vector that bring SARS-CoV-2 into the nursing home.^1, 2, 5^ Identifying asymptomatic and pre-symptomatic carriers has proven challenging, especially before widespread access to testing, and contributed to the risk of transmission in this setting.^6^ Second, most nursing homes are densely populated, often with two (or more) residents sharing a room. This makes it challenging to distance and isolate and increases exposure to SARVS-CoV-2. Third, many certified nursing assistants, who represent 64% of all nursing home staff, hold multiple customer/client facing jobs, which put them at increased risk of COVID-19 infection and thus, transmission to residents.^2, 7-9^ Fourth, most nursing home residents have multiple comorbidities and are immunocompromised, both risk factors for SARS-CoV-2.^10, 11^

Vaccination is a key component of a multi-layered strategy to mitigate the burden of COVID-19 in nursing homes.^12-14^ However, not all residents and staff are vaccinated or up to date with vaccination and breakthrough infections may occur.^15, 16^ Other infection control strategies to manage COVID-19 may include screening residents and staff, having COVID-19 care units, and implementing non-punitive sick leave policies.^17^Improving indoor air quality has also gained attention as an additional strategy to reduce the transmission of SARS-CoV-2 and mitigate COVID-19.^18, 19^ Intervening on indoor air quality is attractive because it can have a wide impact and when implemented at a structural level (i.e., modifying air handling systems) does not depend on human behavior (e.g., wearing a mask). Strategies vary in difficulty with implementation, degree of human involvement, and cost. Each nursing home is eligible to receive $3,000 in Civil Monetary Penalty Reinvestment Program funding for indoor air quality improvements.

The importance of improving indoor air quality is highlighted by data demonstrating SARS-CoV-2 can be transmitted through ventilation systems.^20, 21^ There are myriad interventions to improve indoor air quality, from natural ventilation (e.g., opening windows and doors) to modifying building air handling systems through filtration, purification, dilution, or disinfection.^19, 22^ However, data on the effectiveness of specific air management interventions in nursing homes on SARS-CoV-2 are limited. Thus, we addressed this gap by taking advantage of a ‘natural experiment’ that was conducted when a multi-facility nursing home corporation installed ultraviolet air purification in 84 facilities. Using an interrupted time series design, we examined trends in COVID-19 related outcomes before and after installation in the facilities’ existing heating, ventilation, and air conditioning (HVAC) systems.

## METHODS

### Study Data and Setting

We obtained installation data (facility name, address, and installation date) for 103 nursing homes in a multi-facility corporation in the southern US that gave permission for their distributor to share facility-level data for this evaluation.

These nursing homes installed ultraviolet air purification in their existing HVAC systems. The air purifiers used a photocatalytic process to generate hydrogen peroxide and were installed between July 27, 2020 and September 10, 2020. Technicians from the distributor installed units to account for each facility’s unique HVAC airflow and organic loading. Specific features about the individual nursing homes’ HVAC systems were not available to the research team.

We were able to link 84 of the 103 facilities with the Nursing Home COVID-19 Public Health File, which includes data reported by nursing homes to the CDC’s National Healthcare Safety Network Long-Term Care Facility COVID-19 Module: Surveillance Reporting Pathways and COVID-19 Vaccinations.^4^ Since the week of May 31, 2020, Medicare & Medicaid-certified facilities have submitted weekly COVID-19-related metrics, including the number of residents with a new confirmed COVID-19 case. We also linked nursing homes to data from LTCFocus.org, a publicly-available Brown University database that provides contextual information on nursing home characteristics (e.g., age of residents and number of beds).

We determined the county that each nursing home was located using the address information in the installation data. We then linked county level COVID-19 data from the *New York Times* County COVID-19 Cases,^23^ and outside temperature data with the nursing homes in the distributor file.^24^ The *New York Times* County COVID-19 Cases file provides data on COVID-19 cases and deaths per 100,000 by county. Lastly, temperature data from Spangler et al., contains mean temperature at the county-level on a daily and weekly basis.

### COVID-19-Related Outcomes

We obtained COVID-19 related-outcome data from the Nursing Home COVID-19 Public Health File. Outcomes included weekly confirmed COVID-19 cases per 1,000 residents, whether a nursing home had any COVID-19 cases in the week (0 = no cases; 1= ≥1 case), and whether a nursing home had any COVID-19 deaths in the week (0 = no; 1 = yes).

### Covariates and Contextual Variables

Prior research demonstrates that COVID-19 predominantly enters nursing homes via staff, visitors, and other outside guests.^1, 5^ Therefore, the dynamics of COVID-19 in the surrounding community has a large effect on infections in the nursing home. We controlled for county level time-varying confounders related to community spread of COVID-19. Confounders included weekly COVID-19 cases in the county per 100,000, COVID-19 deaths in the county per 100,000 and mean weekly heat index in the county.

We used data from LTCFocus.org to provide context to the characteristics of the nursing homes in the sample. We did not control for these contextual variables because they are time-invariant and thus cannot confound our associations.

### Statistical Analysis

We described the time-invariant characteristics of the 84 nursing homes. We then used a single group interrupted time series design to compare the trend and immediate change in the outcome in the period before (i.e., pre period) and after (i.e., post period) air purifiers were installed.^25^ We centered time for each nursing home on the week of the air purifier installation. All nursing homes in the analytical file had eight weeks of pre- and eight weeks of post-installation data, which spanned from September 31, 2020 to December 27, 2020. Vaccination began December 18, 2020 and did not confound our analyses.^26^ We used ordinary least squares (OLS) regression to estimate changes in continuous and dichotomous outcomes (i.e., linear probability model). We tested the inclusion of a squared term for time to account for non-linear trajectories, but this did not improve model fit. We clustered standard errors at the nursing home level and accounted for autocorrelation.^27-29^

We conducted all statistical analyses using Stata 16 (College Station, Texas).

## RESULTS

On average, the 84 nursing homes had 108 beds (Standard Deviation or SD: 36.0) and residents were 78.3 years of age (SD: 6.2) (Table 1). The majority of nursing homes were located in Georgia (60.5%) and August 19, 2020 was the median air purifier installation date (range: April 2020 to October 2020).

**Table 1.** Characteristics of Nursing Homes in Pilot Evaluation

|  | Nursing Homes (N = 81) |
| --- | --- |
| **Contextual Measures** |  |
| Location of Nursing Home, n (%) |  |
| Florida | 3 (3.7) |
| Georgia | 49 (60.5) |
| North Carolina | 17 (21.0) |
| South Carolina | 12 (14.8) |
| Percent of female in nursing home, mean (SD) | 65.05 (13.61) |
| Prevalence Black, mean (SD) | 31.50 (24.33) |
| Prevalence White, mean (SD) | 64.53 (24.72) |
| Prevalence Hispanic, mean (SD) | 0.88 (1.28) |
| Age, mean (SD) | 78.27 (6.16) |
| Percentage Admitted from Community, mean (SD) | 5.81 (9.02) |
| **Prior to Intervention, Mean (SD)** |  |
| Weekly COVID-19 cases in the community per 100,000 | 25.0 (14.7) |
| Weekly COVID-19 deaths in the community per 100,000 | 0.4 (0.6) |
| Mean heat index [°C] | 28.4 (2.5) |
| Weekly COVID-19 cases in the nursing home per 1,000 residents | 14.3 (50.8) |
| Proportion of nursing homes with ≥1 COVID-19 case per week (%) | 21.5% |
| Proportion of nursing homes with ≥1 COVID-19 death per week (%) | 8.8% |
| **Post Intervention, Mean (SD)** |  |
| Weekly COVID-19 Cases in the Community per 100,000 | 18.5 (12.4) |
| Weekly COVID-19 Deaths in the Community per 100,000 | 0.7 (0.8) |
| Weekly mean heat index [°C] | 22.4 (11.0) |
| Weekly COVID-19 cases in nursing home per 1,000 residents | 9.3 (46.8) |
| Proportion of nursing homes with ≥1 COVID-19 case per week (%) | 9.6% |
| Proportion of nursing homes with ≥1 COVID-19 death per week (%) | 1.8% |

Figure 1a shows the distribution of COVID-19 cases in the pre and post periods and Figure 1b shows the trend in weekly COVID-19 cases per 1,000 residents in the pre and post periods. In the eight weeks before the air purifiers were installed, the nursing homes had, on average, 14.3 COVID-19 cases per 1,000 residents (SD: 50.8). In the pre period, the number of COVID-19 cases increased by 0.60 (95% Confidence Interval or CI: -1.47, 2.67) per 1,000 residents per week (Table 2). In the post period, there was an immediate decrease in COVID-19 cases (−3.10 cases per 1,000 residents, 95%CI: - 15.75, 9.54), and the number of cases continued to decline on a weekly basis (1.09 cases per week, 95%CI: -2.21, 0.03). This led to 1.69 (95%CI: -4.32, 0.95) fewer COVID-19 cases per 1,000 residents per week in the post compared to pre period.

**Table 2.** Interrupted Time Series Coefficients and 95% Confidence Intervals

|  | Weekly COVID-19 Cases<br>per 1,000 Residents | Any COVID-19 Case per<br>Week (0 = no; 1 = yes) | Any COVID-19 Death per<br>Week (0 = no; 1 = yes) |
| --- | --- | --- | --- |
| Time (pre intervention trend) | 0.60 [-1.47, 2.67] | 0.00 [-0.01, 0.02] | -0.010 [-0.02, 0.00] |
| Treatment Indicator (post intervention intercept) | -3.10 [-15.75, 9.54] | -0.03 [-0.11, 0.06] | -0.01 [-0.07, 0.05] |
| Time * Treatment Interaction | -1.69 [-4.32, 0.95] | -0.02 [-0.04, 0.00] | 0.01 [-0.01, 0.02] |
| Cases per 100k in County | 0.52 [0.17, 0.87] | 0.00 [0.00, 0.01] | 0.00 [0.00, 0.00] |
| Deaths per 100k in County | 0.93 [-2.65, 4.50] | 0.00 [-0.03, 0.04] | 0.00 [-0.02, 0.02] |
| Mean Heat Index | -0.50 [-1.42, 0.42] | 0.00 [-0.01, 0.01] | 0.00 [0.00, 0.01] |
| Intercept | 13.0 [-9.35, 35.42] | 0.13 [-0.08, 0.33] | 0.05 [-0.06, 0.20] |

**Figure 1a.**
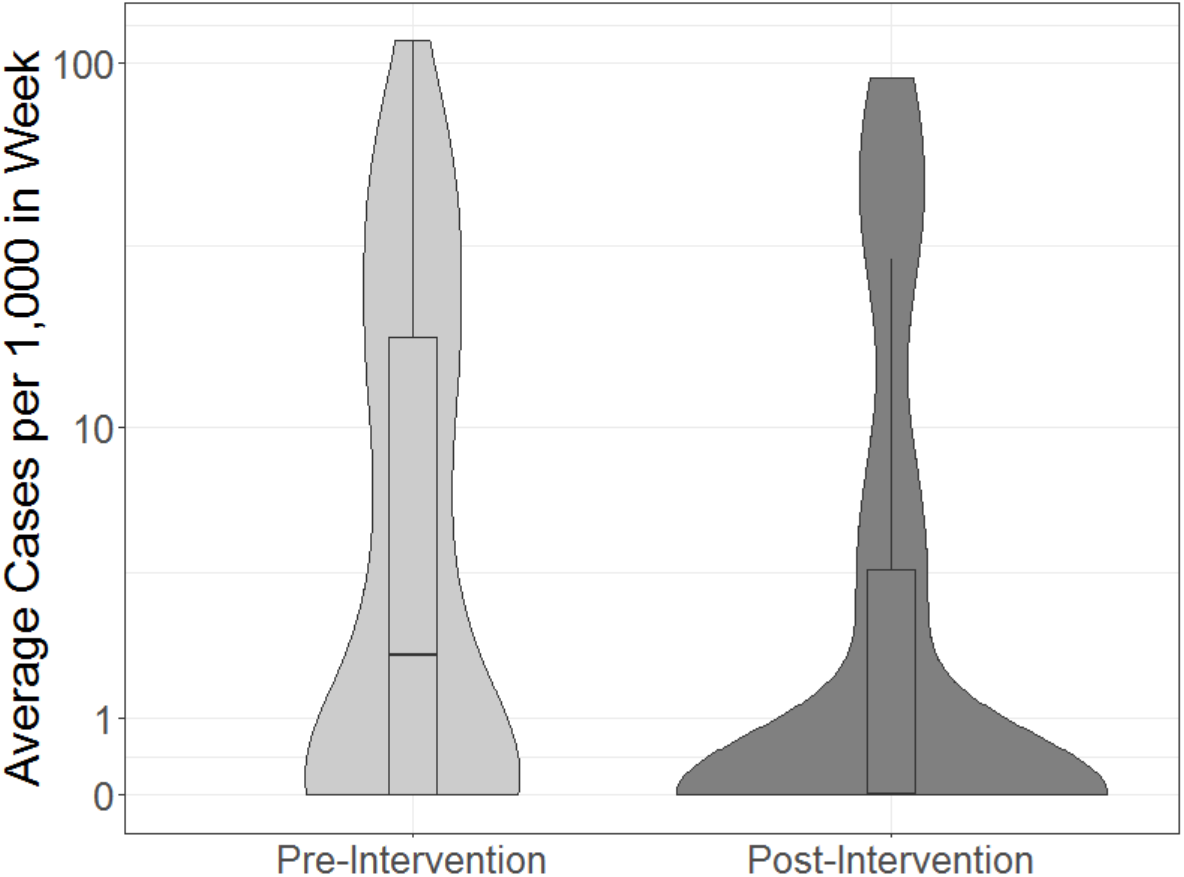
Distribution of Nursing Homes’ Weekly Resident COVID-19 Cases, Pre and Post Periods

**Figure 1b.**
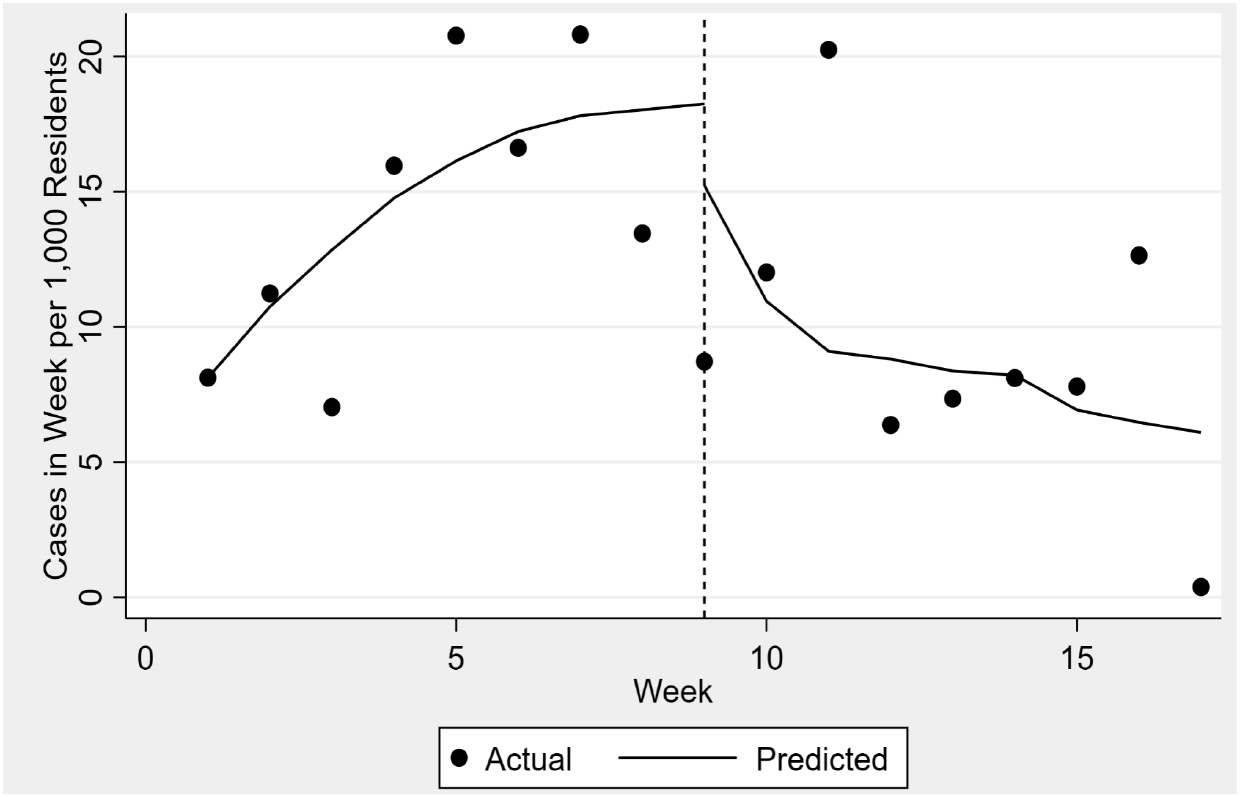
Weekly Cases Per 1,000 Nursing Home Residents Footnote: Dashed vertical line at week 9 separates the pre and post periods.

During the pre period, 21.4% of nursing homes had at least one COVID-19 case per week (Figure 2a). In the first week of the post period, the probability of a nursing home having any COVID-19 case decreased by 0.03 (95%CI: -0.11, 0.06) (Table 2 and Figure 2b). In the post period, the weekly probability of a nursing home having a COVID-19 case decreased by 0.02 (95%CI: -0.04, -0.01). This resulted in a 0.02 decrease in the probability of a nursing home having a COVID-19 cases per week in the post compared to pre period (95%CI: -0.04, 0.00).

**Figure 2a.**
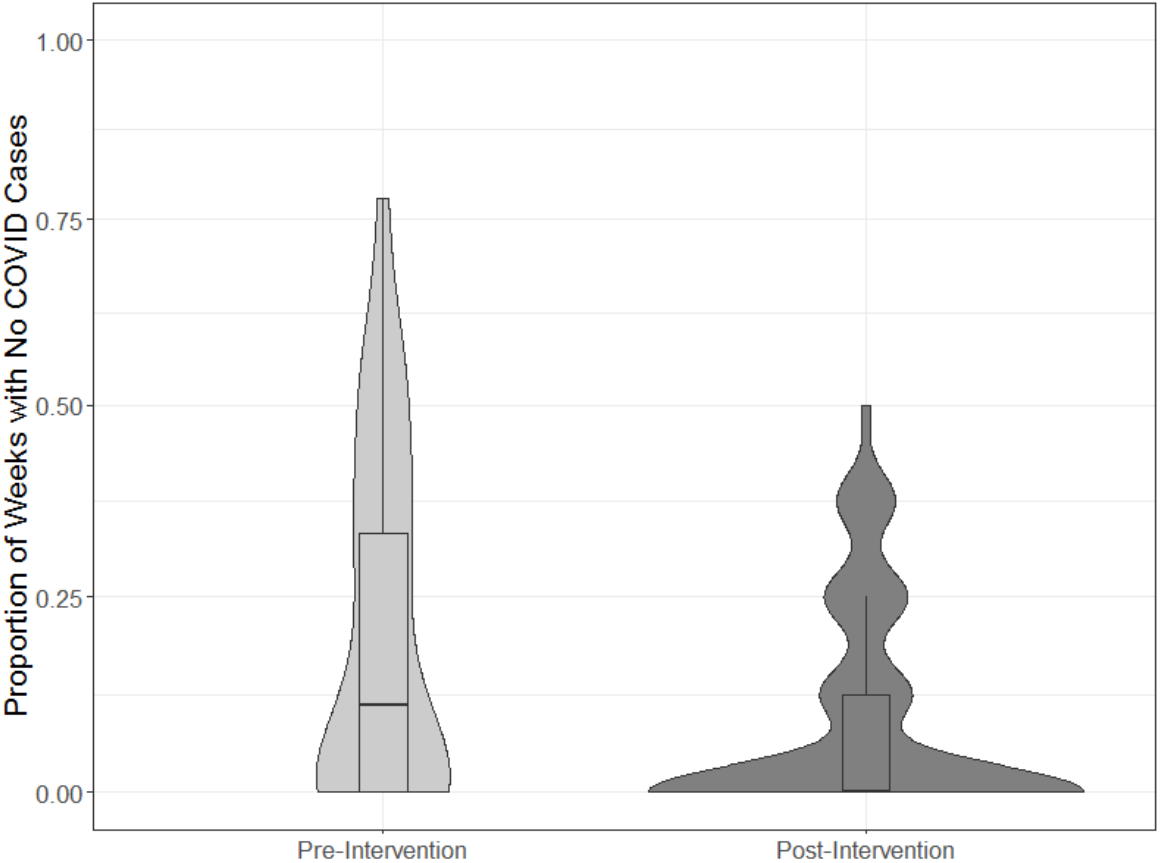
Distribution of Nursing Homes with ≥1 COVID-19 Case in the Pre and Post Period

**Figure 2b.**
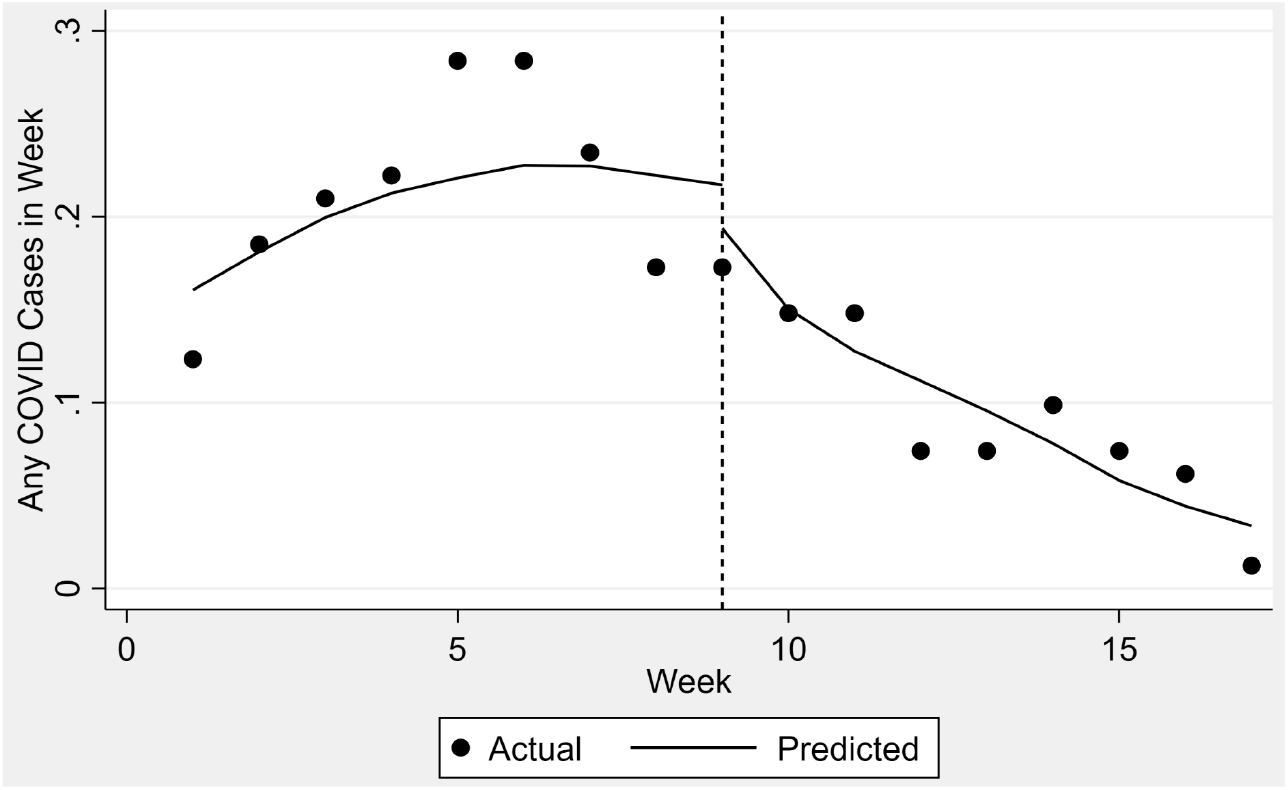
Proportion of Nursing Homes with ≥1 COVID-19 Case Per WeekFootnote: Dashed vertical line at week 9 separates the pre and post period.

Finally, during the pre period, 8.3% of nursing homes had at least one COVID-19 death (Figure 3a). In the pre period there was a decline in the weekly probability of a nursing home reporting a COVID-19 death (Figure 3b). In the first week of the post period, the probability of a nursing home reporting a COVID-19 death decreased by 0.01 percentage points (95%CI: -0.07, 0.05) (Table 2). Finally, in the post period the probability of a nursing home reporting a COVID-19 death per week remained stable, and there was a small increase in the probability of a nursing home reporting a COVID-19 death in the post compared to pre period.

**Figure 3a.**
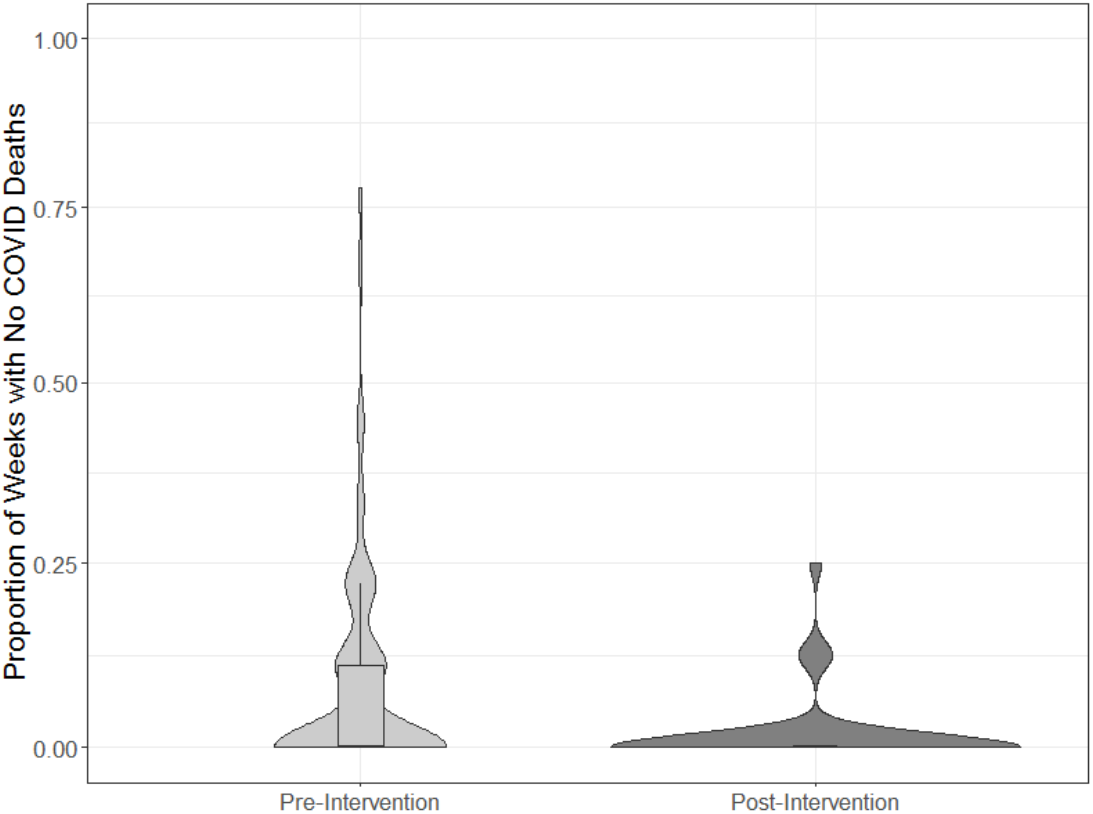
Distribution of Nursing Homes with ≥1 Death, Pre and Post Period

**Figure 3b.**
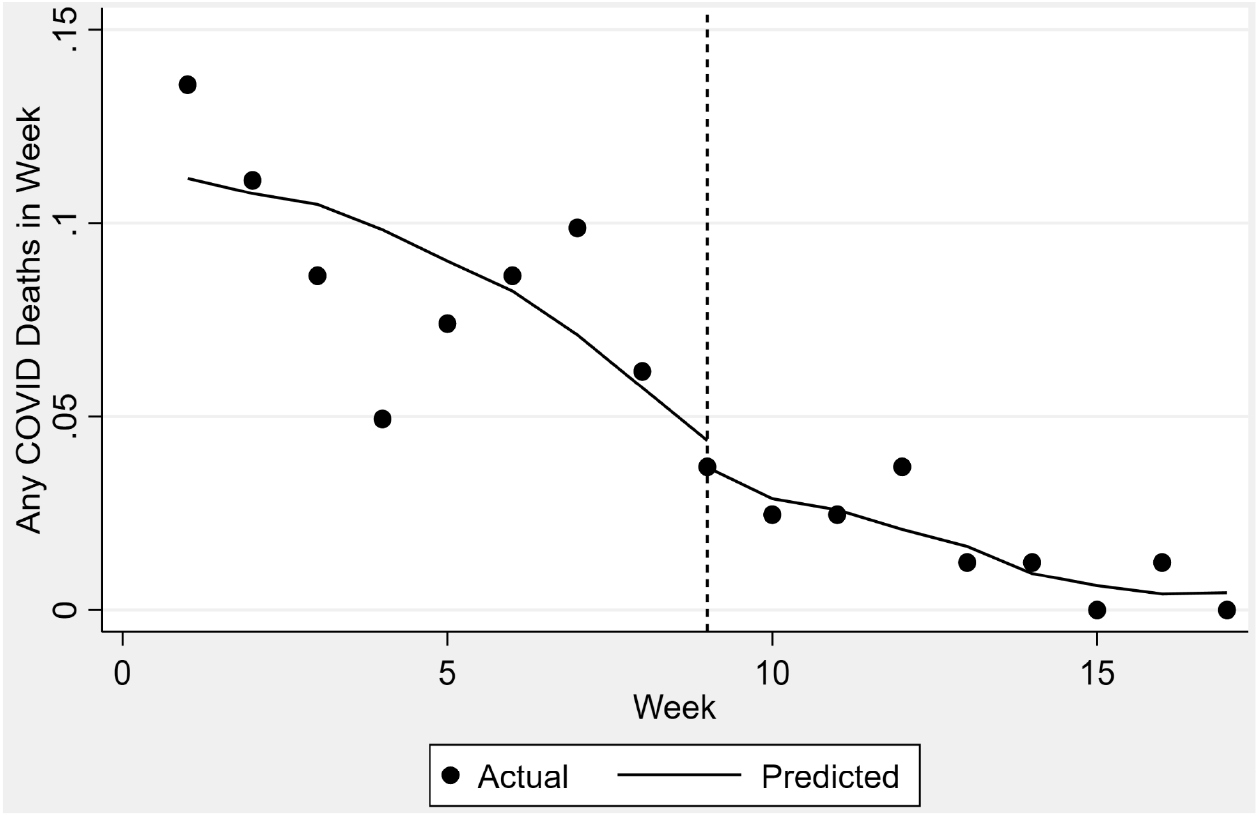
Proportion of Nursing Homes with ≥1 COVID-19 Death Per WeekFootnote: Dashed vertical line at week 9 separates the pre and post period.

## Discussion

Our findings indicate that air purification may be an additional preventative measure for nursing home policymakers and decision makers to consider. We found that COVID-19 cases per 1,000 nursing home residents and the probability of having any COVID-19 case decreased following the installation of ultraviolet air purification in the HVAC systems of 84 nursing homes. Although the effect estimates were imprecise (i.e., wide confidence intervals) and not statistically significant at p<0.05, our pilot evaluation provides the first evidence supporting a potential association between air purification and improved COVID-19 outcomes in this setting, underscoring both the potential and the need for more rigorous research to establish effectiveness.

Improving indoor air quality has gained attention as a strategy to mitigate the risk of SARS-CoV-2 transmission and COVID-19 infection as other layered mitigation strategies have relaxed following widespread vaccination.^19, 30, 31^ For example, a study conducted in 169 Georgia schools found that COVID-19 incidence was lower in schools that improved ventilation compared to those that did not.^32^ Yet, there is wide variation in approaches to improve indoor air quality.^19^ Low-cost interventions, such as opening windows, installing fans, and maintaining HVAC systems, require person-time and oversight.^33^ High-cost interventions, such as modifying HVAC systems, may have a larger impact and require less person-time, but be expensive for nursing homes, which operate on thin margins, to widely adopt.

While COVID-19 cases per 1,000 nursing home residents and the probability of having any COVID-19 case did decrease following installation, we found minimal evidence of an association between installation and the proportion of nursing homes that reported any resident died of COVID-19. However, there was a decline in the pre-intervention COVID-19 cases and the small number of deaths may have limited our ability to identify an association. Larger studies are needed to assess improvements in outcomes following installation of these ultraviolet air purifiers and to examine the comparative effectiveness of different indoor air management strategies. The ultraviolet air purification system installed by the nursing homes in our pilot study is only one type of air filtration and purification system among the many that have been used during the SARS-CoV-2 pandemic. These include both purification and filtration systems that may have variable effectiveness for removing or inactivating airborne SARS-CoV-2 virus.

Importantly, system-level indoor air quality interventions implemented to mitigate SARS-CoV-2 transmission in nursing homes may have positive spillover effects, such as reducing other respiratory pathogens (e.g., influenza).^34^ Residents spend the majority of their time indoors^35^ and there is a growing body of evidence that demonstrates air quality in nursing homes may be suboptimal.^35-37^ Residents, who often have complex comorbidities, may be more vulnerable to adverse health effects of indoor particulates and pollutants.^34^ Future research is needed to assess the impact of improved indoor air quality on other respiratory illnesses and health outcomes.

### Limitations and Strengths

Our pilot evaluation has several important limitations. First, we compared trends in outcomes among facilities in a single multi-facility corporation. There is large variation between nursing homes (e.g., organizational characteristics, physical structure, and resident characteristics), so our findings may not generalize to a broader set of nursing homes. Second, we only observed the date air purifiers were installed. We do not know whether the air purifiers functioned as designed nor do we know what other indoor air quality or other mitigation measures the nursing home staff implemented during the same time period. A strength of our design, however, is that we control for time-invariant confounders, so only time varying confounders pose a threat to the internal validity of our study. Fourth, we did not have detailed information about each nursing home’s HVAC system, although this is unlikely to affect our results given that it is time invariant. Moreover, our effect estimates help answer the question of installing air purifers on COVID-19 outcomes. We do not answer questions about the aspects of nursing homes that impact efficacy or what type of air interventions are most effective work. Fifth, nursing homes self-report their weekly COVID-19-related outcomes to the CDC. During the time period of the study there may have been variation in staff’s ability to diagnose and manage COVID-19.^38, 39^ Sixth, intervening on indoor air quality may both lessen risk of exposure and reduce dose of exposure, which may be linked to severity of illness.

Finally, we evaluated outcomes that occurred between May 2020 and December 2020. This is a period before vaccines were available and where there were significant precautions in place to prevent mortality.^40^ Furthermore, the time period of analysis includes summer 2020, a time when community transmission declined.^41, 42^ It will be important to understand the effect of indoor air quality interventions, such as air purification, in the context of vaccines.

## Conclusion

Intervening on indoor air quality has great potential to have a large impact on SARS-CoV-2 mitigation in nursing homes with minimal human involvement. Minimizing the concentrations of airborne SARS-CoV-2 is particularly important in the context of nursing homes where residents are high risk, have limited mobility, are inside for most of the day, and are dependent on staff for basic activities of daily living. In our pilot evaluation, weekly COVID-19 cases per 1,000 residents decreased among nursing homes after installation of an air purifier compared to before the installation. Although the effect was large, it was imprecisely estimated and not statistically significant and there was no evidence of an association between installation and the proportion of nursing homes reporting a COVID-19 death per week. Although there are robust data from schools and other indoor settings demonstrating improvements in COVID-19 and other health outcomes, we recommend a larger study with diverse nursing homes and a stronger experimental design to assess the causal effect of indoor air interventions, such as air purification, on COVID-19 in this setting. Setting-specific data can guide policy and practice, including strategies to fund and maintain indoor air quality improvements.

## Data Availability

Public data used in analyses are directly available from the sources or the authors. We linked public data with private data from a multi-facility nursing home corporation in the southern US that gave permission for the distributor (Smart Air Care) of the ultraviolet air purification system that they installed to share facility-level data for this evaluation.

